# Internal Dose Benchmarking of Acrylamide Exposure and Peripheral Neuropathy Risk: A National Population-Based Validation Study

**DOI:** 10.64898/2026.08.10.26360101

**Authors:** Khaled Hamed, Ruweida M. Bundid, Maha A. Sayah, Rawan S. M. Taha, Nahid Nuri, Mohamed Gamal

## Abstract

**Background:** Acrylamide, a neurotoxicant in heated foods and smoke, is linked to occupational neuropathy, but evidence regarding chronic, low-level population exposure remains limited. We evaluated the association between acrylamide exposure biomarkers and peripheral neuropathy among U.S. adults.

**Methods:** A total of 2,266 NHANES 2003–2004 participants (age ≥40) were analyzed. Exposure was assessed via hemoglobin adducts (HbAA/HbGA); neuropathy via monofilament testing (≥1 site). Survey-weighted logistic regression models adjusted for confounders. Sensitivity analyses included cubic splines, diabetes stratification, and multiple imputation.

**Results:** Neuropathy prevalence was 15.5%. In adjusted models, neither adduct was associated with neuropathy (HbAA OR: 0.98, 95% CI: 0.82–1.17; HbGA OR: 0.91, 95% CI: 0.77–1.08). No dose-response gradient was observed. Expected risk factors (age, diabetes) showed strong associations, validating model sensitivity. The null result remained robust across sensitivity analyses, including a stricter outcome definition and multiple imputation (pooled OR: 0.97, 95% CI: 0.83–1.14).

**Conclusions:** Acrylamide adducts were not associated with peripheral neuropathy in this national sample. General population levels (∼55–70 pmol/g) lie well below established occupational no-observed-adverse-effect levels (∼510 pmol/g) and clinical neuropathy thresholds (∼6,000 pmol/g), providing a mechanistically coherent explanation for this null result.

## 1. Introduction

Acrylamide is a low-molecular-weight, water-soluble α,β-unsaturated carbonyl compound historically used in industrial polyacrylamide manufacture, wastewater treatment, and construction grouting (Rifai & Saleh, 2020). Public health interest expanded markedly after 2002, when it was discovered to form in carbohydrate-rich foods heated above 120 °C through the Maillard reaction between asparagine and reducing sugars (Lineback et al., 2012; Pedreschi et al., 2014;Törös et al., 2026). Major dietary contributors include coffee, fried potato products, and cereals; additionally, tobacco smoke remains a significant non-dietary source (Halford & Raffan, 2019). Consequently, the general population is exposed chronically to low doses, contrasting sharply with the high-level intermittent exposures encountered in occupational settings (World Health Organization, 2002).

Acrylamide is a well-characterized neurotoxicant and a Group 2A “probable human carcinogen” (IARC) (Hogervorst et al., 2010; Klaunig, 2008). Following absorption, acrylamide is handled by two competing pathways: detoxifying conjugation with glutathione, catalysed in part by glutathione S-transferases, and cytochrome P450 2E1–mediated oxidation to glycidamide, a reactive epoxide that is considered the proximate agent of much of the compound’s genotoxicity and carcinogenicity (Behrens et al., 2019; Calleman, 1996; Fennell & Friedman, 2005; Kraus et al., 2013). Both the parent compound and its metabolite are electrophiles that form covalent adducts at nucleophilic sites on proteins and DNA, and both are neurotoxic (Costa et al., 1995; Doerge et al., 2008; Koyama et al., 2006).

Mechanistically, acrylamide induces a prototypical central–peripheral distal axonopathy. Neurotoxicity is attributed to Michael-type addition to cysteine thiol groups on presynaptic and axonal proteins, including components of the neurotransmitter-release and fast-axonal-transport machinery, with consequent impairment of vesicle trafficking, progressive nerve-terminal degeneration, and—at higher cumulative exposures—retrograde axonal degeneration (Harris et al., 1994; LoPachin & DeCaprio, 2005; Valentine et al., 1997). These neurological, morphological, and molecular endpoints have been extensively characterized in various animal models (LoPachin, 2005).. Additional contributions from neurofilament dysregulation, oxidative stress, and disturbed calcium/calmodulin-dependent signalling have been described, and glycidamide adds a further genotoxic and neurotoxic dimension (Reagan et al., 1994; Zhao et al., 2022). A pivotal feature of acrylamide neurotoxicity is dose-rate dependence: low, protracted exposure produces qualitatively milder lesions than the same total dose delivered rapidly(Crofton et al., 1996; LoPachin et al., 2000). This dose-rate dependence is central to interpreting risk at the low, steady internal doses that characterize dietary exposure.

Most human evidence of acrylamide-related peripheral neuropathy derives from occupational cohorts exposed at levels far exceeding dietary intake. Symptoms observed in factory and construction workers are generally mild and reversible upon cessation of exposure (Bin-Jumah et al., 2021; Goffeng et al., 2008; Kim et al., 2000; Li et al., 2024).Crucially, these studies established internal dose thresholds using hemoglobin adducts. Dose-response modeling among tunnel workers placed the emergence of symptoms at approximately 1,000 pmol/g (Hagmar et al., 2001). Independent estimates derived a no-observed-adverse-effect level (NOAEL) of ∼2,000 pmol/g and a lowest-observed-adverse-effect level (LOAEL) near 6,000 pmol/g for clinical neuropathy (Bergmark, 1997; Calleman et al., 1994). These occupational benchmarks provide an indispensable yardstick against which general-population internal doses can be judged, and they frame acrylamide neuropathy as a threshold phenomenon rather than a linear, no-threshold hazard (Albers & Berent, 2005; Berger & Pulley, 2014).

Accurate exposure classification is the principal obstacle to studying dietary acrylamide. Food-frequency questionnaires capture intake imperfectly, whereas hemoglobin adducts of acrylamide (HbAA) and glycidamide (HbGA) integrate exposure over the 120-day erythrocyte lifespan (Bergmark et al., 1993; DeCaprio, 1997; Lockridge, 2023; Meyer & Bechtold, 1996; Ogawa et al., 2006; Pérez et al., 1999). For this reason, hemoglobin adducts are widely regarded as the reference biomarker of internal acrylamide dose in both occupational and general populations. In the 2003–2004 National Health and Nutrition Examination Survey (NHANES), the CDC quantified these adducts using validated mass spectrometry (Vesper et al., 2006), creating a unique opportunity to link internal dose to health outcomes at a national scale.

Despite strong biological plausibility, whether chronic low-level exposure produces subclinical neuropathy in the general population has not been tested using biomarker-confirmed doses and objective neurological outcomes. Prior NHANES analyses have concentrated on metabolic endpoints or central cognition rather than peripheral nerve function (Hung et al., 2021; Lin et al., 2009; Liu et al., 2017; Xu et al., 2025; Yin et al., 2022; Yu et al., 2023). The same adduct-biomarker paradigm has been applied to related electrophiles, such as ethylene oxide, further establishing its analytic feasibility within NHANES (Zhou et al., 2024). No study, to our knowledge, has evaluated whether HbAA or HbGA is associated with objectively assessed peripheral neuropathy in a nationally representative sample.

We used NHANES 2003–2004—the only survey cycle pairing adduct measurements with standardized monofilament testing—to evaluate whether higher internal-dose biomarkers are associated with increased odds of peripheral neuropathy among U.S. adults aged ≥ 40 years. By combining objective biomarkers for both exposure and outcome, this analysis addresses a previously untested question at the intersection of environmental neurotoxicology and neuroepidemiology..

## 2. Methods

### 2.1. Study design and participants

Data were extracted from the 2003–2004 National Health and Nutrition Examination Survey (NHANES), a nationally representative, multistage probability survey of the U.S. civilian noninstitutionalized population (CDC, 2014). NHANES 2003–2004 was the only cycle to concurrently measure hemoglobin adducts and perform lower-extremity monofilament examinations. Because neurological testing was restricted to older participants, our target population comprised adults aged ≥ 40 years. Of 10,122 participants, 2,266 provided complete exposure and outcome data; 2,049 had complete covariate data for fully adjusted models (Fig. 1)

**Fig. 1.**
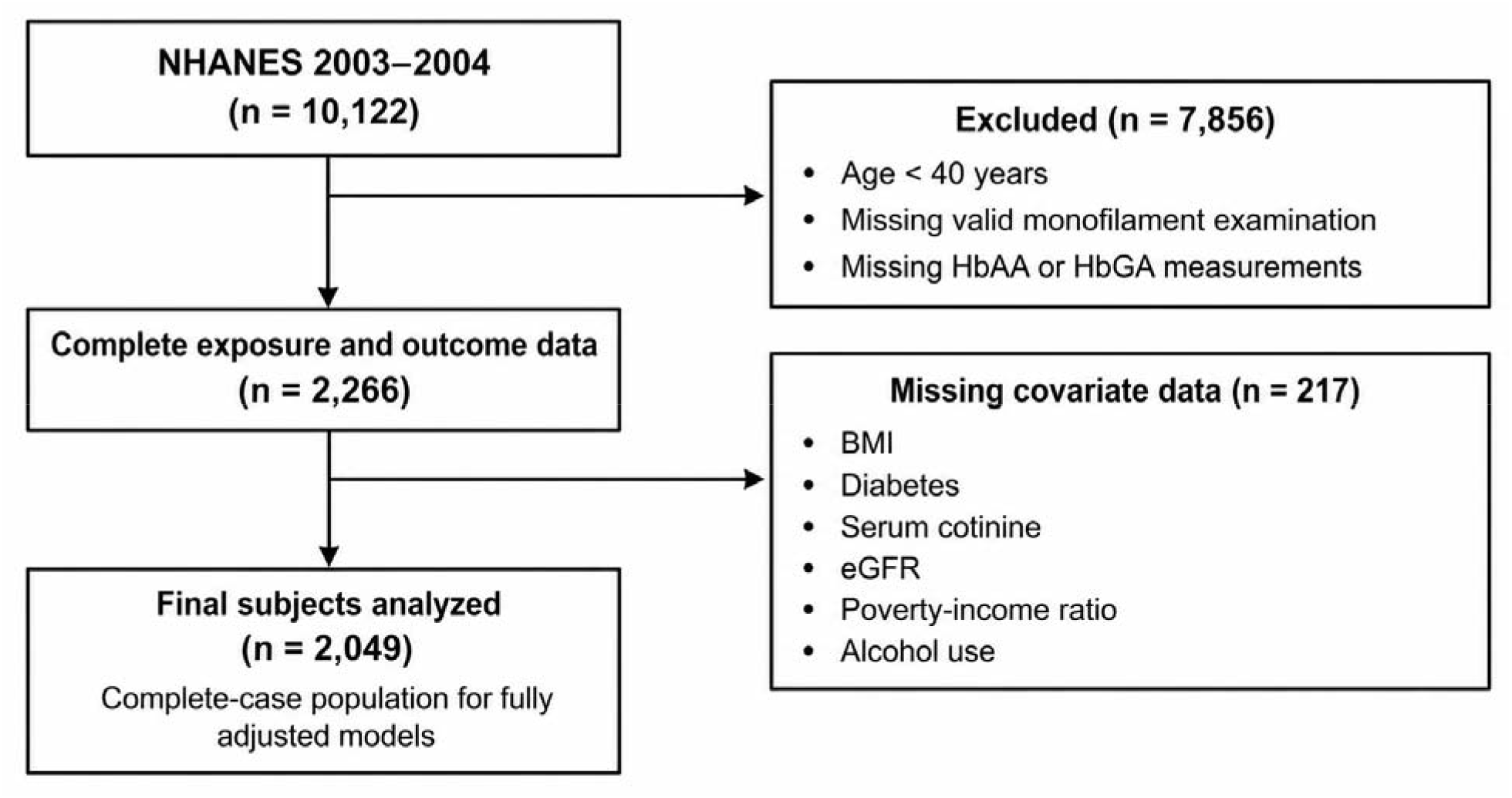
Flow chart of the selection of eligible subjects

### 2.2. Demographics and covariates

Covariates included age, sex, race/ethnicity, family poverty-income ratio (PIR), body-mass index (BMI), estimated glomerular filtration rate (eGFR), alcohol use, and serum cotinine. Diabetes was defined via a composite criterion: self-reported diagnosis, glycated hemoglobin (HbA1c) ≥ 6.5%, or fasting plasma glucose ≥ 126 mg/dL. eGFR was calculated using the 2021 race-free CKD-EPI creatinine equation (Inker et al., 2021). Serum cotinine (ng/mL), a biomarker for active and passive smoke exposure, was measured by isotope-dilution HPLC/MS/MS and modeled as a continuous covariate.

### 2.3. HbAA and HbGA measurement

Hemoglobin adducts (HbAA/HbGA) were quantified in red blood cells using HPLC/tandem mass spectrometry (HPLC/MS/MS) targeting the N-terminal valine (Vesper et al., 2010). Concentrations were expressed as pmol/g hemoglobin. Due to high metabolic correlation, adducts were entered into separate single-biomarker models. Each biomarker was z-score standardized and additionally parameterized as quartiles, natural logarithms, and restricted cubic splines.

### 2.4. Peripheral neuropathy assessment

Neuropathy was ascertained via standardized Semmes–Weinstein 5.07 (10-g) monofilament testing at three sites on the plantar surface of each foot. Sites not perceived by the participant were classified as insensate. The primary outcome was “any peripheral neuropathy” (≥1 insensate site), with a stricter definition (≥2 sites) utilized for secondary analysis.

### 2.5. Statistical analysis

Analyses accounted for the NHANES complex survey design using examination sample weights (WTMEC2YR), stratification (SDMVSTRA), and primary sampling units (SDMVPSU), with variances estimated via Taylor-series linearization. The realized design provided 15 degrees of freedom for design-based tests. Weighted characteristics were compared by neuropathy status using design-based t-tests and Rao–Scott chi-square tests. Adduct interdependence was assessed via correlation coefficients and variance inflation factors.

Survey-weighted quasibinomial logistic regression estimated odds ratios (ORs) and 95% confidence intervals (CIs) for peripheral neuropathy. For each biomarker, we constructed three nested models per 1-SD increment:

**Model 1**: Unadjusted.

**Model 2**: Adjusted for age, sex, and race/ethnicity.

**Model 3**: Fully adjusted for demographics plus BMI, diabetes, serum cotinine, eGFR, PIR, and alcohol use.

Exposure–response relationships were evaluated via adduct quartiles and restricted cubic splines (Desquilbet and Mariotti, 2010). Potential effect modification by diabetes was assessed using multiplicative interaction terms and stratified analysis.

Robustness was verified through pre-specified sensitivity analyses: restricting the sample to non-smokers (low cotinine) and non-diabetic participants, applying a stricter outcome definition (≥2 sites), utilizing log-transformed exposures, and substituting the HbGA/HbAA ratio as a proxy for metabolic activation. Missing covariate data were addressed via multiple imputation by chained equations (10 imputations). To contextualize null findings, we determined the minimum detectable OR at 80% power and calculated E-values to characterize the strength of unmeasured confounding required to negate the observed estimates (VanderWeele and Ding, 2017). The analytical pipeline was cross-validated in R (v4.6.0) and Python (v3.12.13), with the complete computational notebook available as an open-source supplement (Hamed et al., 2026)..

## 3. Results

### 3.1. Study population characteristics

The 2,266 participants aged ≥40 years represented approximately 95.8 million U.S. adults (weighted mean age 56.7 years). Peripheral neuropathy, defined by at least one insensate site on monofilament testing, was present in 467 participants (weighted prevalence 15.5%), and 180 (5.4%) had two or more insensate sites. As shown in Table 1, participants with neuropathy were significantly older than those without (weighted means 63.0 vs 55.5 years), more often male (68.2% vs 43.4%), and more often diabetic (32.0% vs 12.0%), with higher glycated hemoglobin (6.07% vs 5.60%), lower estimated glomerular filtration rate (81.6 vs 88.6 mL/min/1.73 m²), and a higher prevalence of hypertension (53.9% vs 40.1%) (all p ≤ 0.001). Current smoking and alcohol use did not differ by neuropathy status. Notably, and contrary to the hypothesized direction, hemoglobin adduct concentrations were lower among participants with neuropathy than among those without (HbGA 60.4 vs 69.2 pmol/g, p = 0.029; HbAA 67.9 vs 74.5 pmol/g, p = 0.092).

**Table 1.** Weighted characteristics of the study population by peripheral neuropathy status, NHANES 2003–2004.

| Characteristic | No neuropathy | Neuropathy | p-value |
| --- | --- | --- | --- |
| Age, years | 55.5 | 63.0 | <0.001 |
| Female, % | 56.6 | 31.8 | <0.001 |
| Body-mass index, kg/m<sup>2</sup> | 28.6 | 29.4 | 0.020 |
| Diabetes, % | 12.0 | 32.0 | <0.001 |
| Glycated hemoglobin, % | 5.60 | 6.07 | <0.001 |
| eGFR, mL/min/1.73 m<sup>2</sup> | 88.6 | 81.6 | <0.001 |
| Hypertension, % | 40.1 | 53.9 | 0.001 |
| Current smoker, % | 21.2 | 19.9 | 0.621 |
| Any alcohol use, % | 68.6 | 68.8 | 0.934 |
| Income-to-poverty ratio | 3.24 | 2.94 | 0.002 |
| HbAA, pmol/g | 74.5 | 67.9 | 0.092 |
| HbGA, pmol/g | 69.2 | 60.4 | 0.029 |

Values are survey-weighted means or percentages. *p*-values from design-based *t*-tests (continuous variables) and Rao–Scott chi-square tests (categorical variables). Analytic *n* = 2,266 (467 with neuropathy). eGFR, estimated glomerular filtration rate; HbAA/HbGA, hemoglobin adducts of acrylamide/glycidamide.

### 3.2. HbAA and HbGA levels

The geometric mean (GM) for HbAA was 57.8 pmol/g and 51.8 pmol/g for HbGA, with both adducts exhibiting a pronounced right skew (Table 2). Concentrations were significantly higher in smokers compared to non-smokers (HbAA GM: 125.8 vs. 48.1 pmol/g; HbGA GM: 92.9 vs. 45.1 pmol/g), identifying tobacco smoke as a primary exposure determinant. Due to high inter-adduct correlation (Pearson r=0.84) and elevated variance inflation factors (VIF ≥3.6), single-biomarker models were utilized as the primary analytic approach.

**Table 2.** Hemoglobin adduct concentrations (pmol/g Hb) by age, sex, and smoking status, NHANES 2003–2004.

| Subgroup | HbAA, GM (95% CI) | HbAA, median (IQR) | HbGA, GM (95% CI) | HbGA, median (IQR) |
| --- | --- | --- | --- | --- |
| All | 57.8 (56.5–59.2) | 51.8 (39.8–73.6) | 51.8 (50.4–53.3) | 51.8 (37.7–74.2) |
| **Age** |  |  |  |  |
| 40–64 years | 65.5 (63.4–67.8) | 57.0 (42.4–92.5) | 58.4 (56.2–60.7) | 57.9 (41.5–84.9) |
| ≥65 years | 49.1 (47.7–50.5) | 46.6 (37.6–59.7) | 44.3 (42.6–46.1) | 45.6 (33.5–62.0) |
| **Sex** |  |  |  |  |
| Male | 60.4 (58.3–62.5) | 53.3 (40.3–81.2) | 50.8 (48.7–53.0) | 50.9 (36.0–74.5) |
| Female | 55.4 (53.7–57.1) | 50.2 (39.7–68.8) | 52.8 (50.9–54.9) | 52.7 (39.7–73.8) |
| **Smoking status** |  |  |  |  |
| Non-smoker | 48.1 (47.2–49.0) | 47.1 (38.0–59.2) | 45.1 (43.9–46.4) | 47.5 (35.5–63.5) |
| Current smoker | 125.8 (119.7–132.3) | 129.5 (89.7–179.0) | 92.9 (87.3–98.9) | 97.9 (65.5–139.0) |

*GM, geometric mean; CI, confidence interval; IQR, interquartile range. Concentrations expressed as pmol/g hemoglobin.*

### 3.3. Associations between HbAA and HbGA levels and peripheral neuropathy

Adjusted odds ratios for peripheral neuropathy per one–standard-deviation increment and across quartiles of HbAA and HbGA are presented in Table 3, with covariate estimates in Table 4; the overall progression of these models is visually summarized in Fig. 2. In the fully adjusted model, the odds ratio per 1-SD was 0.98 (95% CI 0.82–1.17) for HbAA and 0.91 (0.77–1.08) for HbGA; neither was statistically significant. The crude HbAA estimate lay below the null (0.89, 0.77– 1.02) but moved to the null after adjustment for age, sex, and race/ethnicity (0.98, 0.86–1.11) and remained there with full adjustment, indicating that the crude inverse association reflected confounding rather than a protective effect. Across HbAA quartiles, odds ratios relative to the lowest quartile were 1.37 (0.90–2.09), 1.17 (0.81–1.69), and 1.27 (0.73–2.21), with no evidence of a monotonic trend (p-trend = 0.47). For HbGA, the second quartile showed a nominally lower odds (0.73, 0.55–0.98), but the third and fourth quartiles were null (0.96 and 0.85) and the trend test was non-significant (p-trend = 0.75), consistent with a chance finding among multiple comparisons. In the same fully adjusted model, established determinants of neuropathy were associated with the outcome in the expected directions—older age (OR 1.05 per year), female sex (OR 0.32), and diabetes (OR 2.62) (Table 4)—supporting the internal validity of the analytic approach and indicating that established neuropathy risk factors were detected as anticipated.

**Figure 2.**
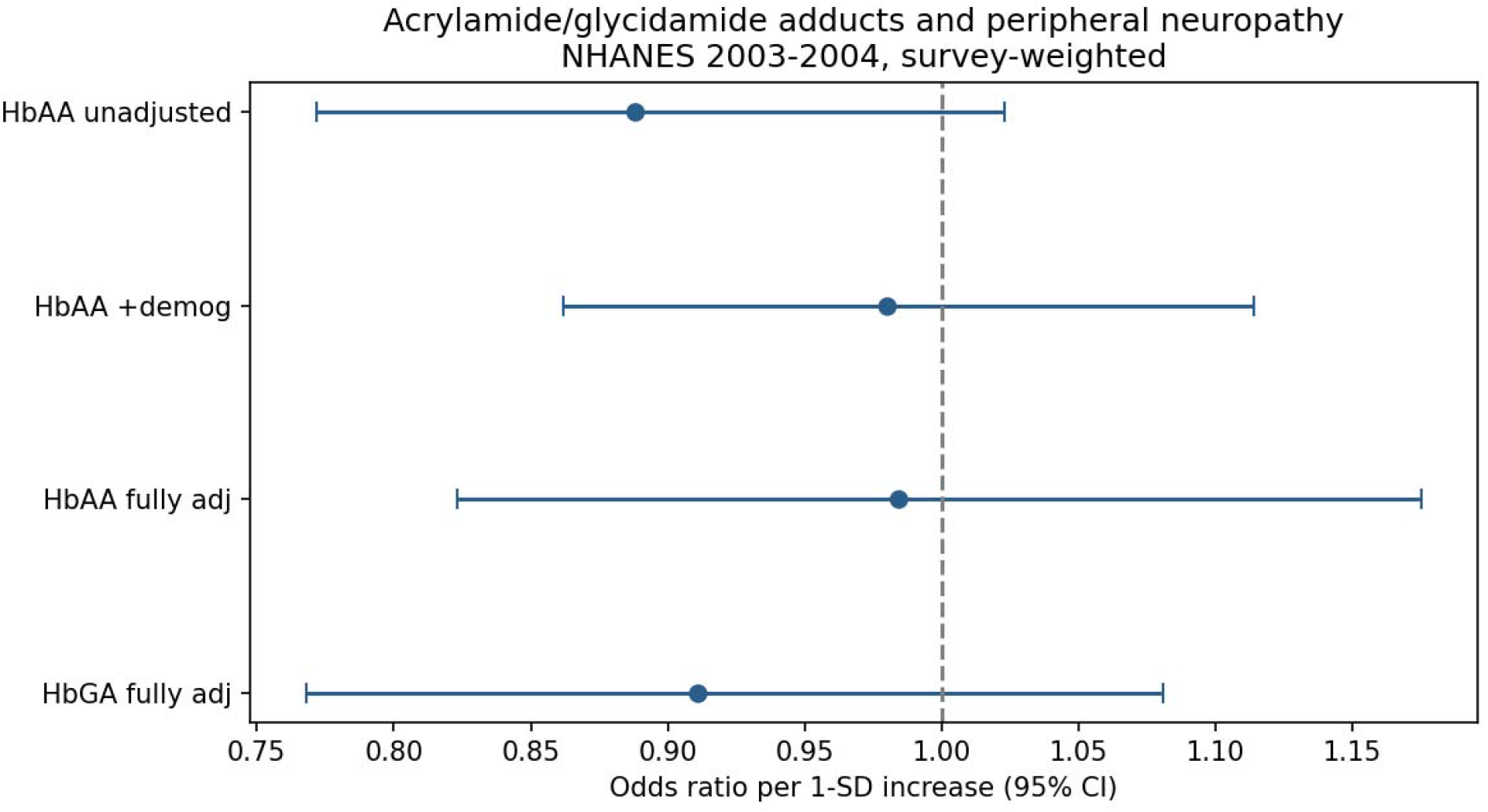
Survey-weighted odds ratios (ORs) and 95% confidence intervals (CIs) for peripheral neuropathy per 1-SD increase in HbAA and HbGA across nested logistic regression models. Model 1 was unadjusted, Model 2 adjusted for age, sex, and race/ethnicity, and Model 3 additionally adjusted for BMI, diabetes, serum cotinine, eGFR, poverty-income ratio, and alcohol use. Neither biomarker was significantly associated with peripheral neuropathy after adjustment.

**Table 3.**
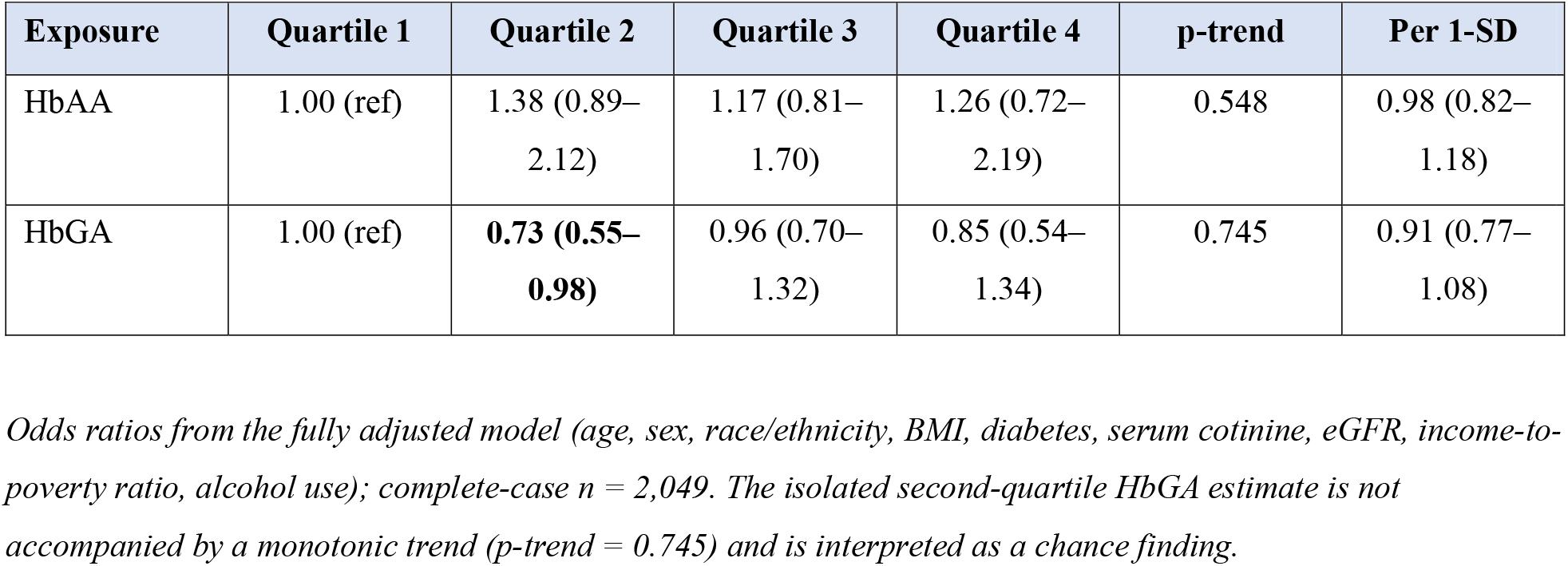
Survey-weighted odds ratios (95% CI) for peripheral neuropathy by quartile and per 1-SD of HbAA and HbGA.

**Table 4.** Covariate (positive-control) associations with peripheral neuropathy in the fully adjusted model.

| Covariate | OR (95% CI) | p-value |
| --- | --- | --- |
| Age (per year) | **1.05 (1.03–1.08)** | **0.015** |
| Female sex | **0.32 (0.19–0.53)** | **0.011** |
| Diabetes | **2.63 (1.13–6.11)** | **0.039** |
| Body-mass index (per kg/m<sup>2</sup>) | 1.03 (1.00–1.06) | 0.181 |
| Income-to-poverty ratio | 0.91 (0.84–0.99) | 0.053 |
| Serum cotinine (per ng/mL) | 1.00 (1.00–1.00) | 0.463 |
| eGFR (per mL/min/1.73 m<sup>2</sup>) | 1.00 (0.99–1.01) | 0.983 |
| Any alcohol use | 1.03 (0.68–1.55) | 0.896 |

*Estimates from the fully adjusted logistic model. Values in boldface denote $p < 0.05$ . These covariate associations* *serve as internal positive controls.*

### 3.4. Dose–response, effect modification, and sensitivity analyses

Restricted cubic spline models provided no evidence of an association for either biomarker overall p = 0.61 for HbAA and 0.61 for HbGA and no departure from linearity (p = 0.43 and 0.51; Fig. 3) As shown by the spline curves, the predicted probability of neuropathy was essentially flat across the exposure range for both adducts. There was no effect modification by diabetes on either the multiplicative scale (interaction p = 0.236) or the additive scale (relative excess risk due to interaction −0.10, 95% CI −1.28 to 1.08), and the HbAA estimate was null within both non-diabetic (OR 0.99, 0.90–1.09) and diabetic (OR 1.14, 0.90–1.46) strata (Fig. S1). The null association was robust across all sensitivity analyses (Table 5), including restriction to participants with low serum cotinine (OR 0.85), restriction to non-diabetic participants (0.99), the more specific ≥2-site outcome (1.15), a log-transformed exposure (1.11), and multiple imputation of missing covariates (0.98, 0.84–1.14). The metabolic-activation ratio (HbGA/HbAA), which was inversely associated with neuropathy in an unweighted analysis, was null under design-based estimation (0.68, 0.43–1.08). On the basis of the observed precision, the study had approximately 80% power to detect an odds ratio of 1.29 per 1-SD; however, smaller associations cannot be excluded.

**Figure 3.**
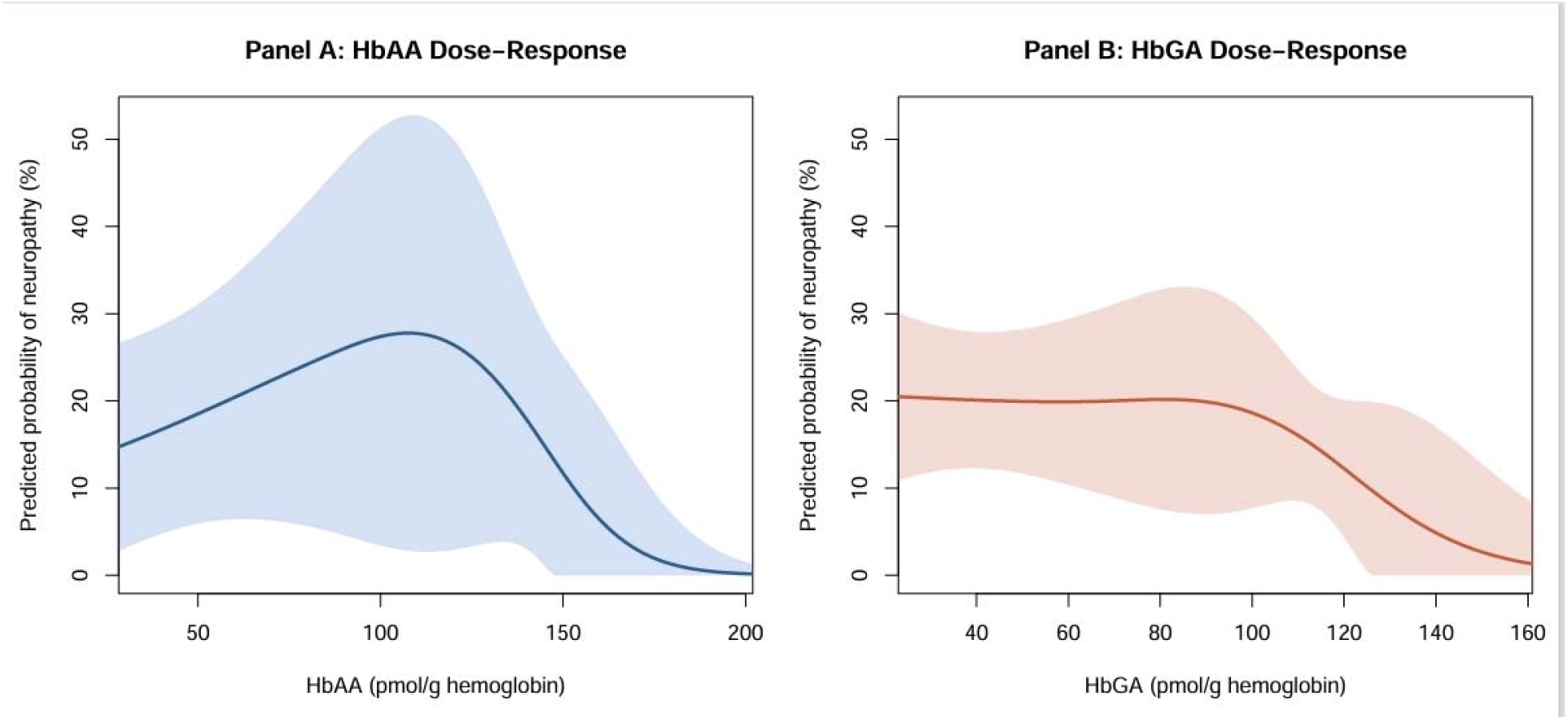
Restricted cubic spline analyses of the association between acrylamide biomarkers and peripheral neuropathy. Panel A shows HbAA and Panel B shows HbGA. Solid lines represent adjusted odds ratios and shaded areas indicate 95% confidence intervals. No significant overall or non-linear dose–response relationship was observed.

**Table 5.** Sensitivity analyses for the association between HbAA and peripheral neuropathy.

| Analysis | n | OR (95% CI)<sup>a</sup> | p-value |
| --- | --- | --- | --- |
| **Primary Continuous Models** |  |  |  |
| HbAA (per 1-SD) | 2049 | 0.98 (0.82–1.17) | 0.858 |
| HbGA (per 1-SD) | 2049 | 0.91 (0.77–1.08) | 0.301 |
| **Diabetes Interaction & Stratification<sup>b</sup>** |  |  |  |
| Multiplicative interaction (HbAA x Diabetes) | 2049 |  | 0.236 |
| Non-diabetic stratum | 1807 | 0.99 (0.90–1.09) | 0.852 |
| Diabetic stratum | 436 | 1.14 (0.90–1.46) | 0.308 |
| Additive interaction (RERI)<sup>c</sup> | 2049 | -0.10 (-1.28–1.08) | — |
| **Robustness & Sensitivity Analyses (HbAA)** |  |  |  |
| Cotinine <10 ng/mL subgroup | 1704 | 0.85 (0.66–1.09) | 0.238 |
| Non-diabetic subgroup | 1807 | 0.99 (0.89–1.09) | 0.790 |
| Stricter outcome ($\geq 2$ sites) | 2049 | 1.17 (0.90–1.51) | 0.251 |
| Log-transformed HbAA | 2049 | 1.10 (0.91–1.34) | 0.356 |
| HbGA/HbAA metabolic-activation ratio | 2049 | 0.69 (0.43–1.08) | 0.141 |
| Multiple imputation (pooled, $m = 10$)<sup>d</sup> | 2266 | 0.97 (0.83–1.14) | 0.732 |

<sup>a</sup> All models are adjusted for age, gender, race/ethnicity, body-mass index, diabetes status, serum cotinine (ln-transformed), GFR, poverty-income ratio, and alcohol use, unless restricted by the subgroup definition.
<sup>b</sup> Dichotomized at the survey-weighted median (52.4 pmol/g Hb).
<sup>c</sup> RERI centered near zero indicates a complete absence of additive interaction.
<sup>d</sup> Missing covariates were multiply imputed using chained equations (10 imputations) and pooled using Rubin's rules to restore the full sample size of 2,266.

## 4. Discussion

### 4.1 Principal findings

In this nationally representative sample of 2,266 U.S. adults representing approximately 95.8 million individuals, neither HbAA nor HbGA was associated with peripheral neuropathy. In fully adjusted models—controlling for demographics, glycemic status, smoking (via questionnaire and serum cotinine), and socioeconomic factors—the odds ratio for HbAA per standard-deviation increase was approximately unity (OR: 0.98; 95% CI: 0.82–1.17); the estimate for HbGA was likewise null. Quartile and restricted cubic-spline analyses revealed no monotonic dose-response or non-linear associations, and mutual adjustment of the two adducts did not unmask hidden signals. This null result remained robust across all pre-specified sensitivity analyses, including multiple imputation, stricter outcome definitions, and stratification by smoking or diabetes status.

Two features establish that this null is informative rather than an artifact of low statistical power or poor validity. First, established risk factors—older age, higher BMI, and, most significantly, diabetes—showed robust, significant associations in the expected directions, confirming the internal validity of the analytical approach. Second, a formal power analysis indicated the study was adequately powered to detect modest odds ratios expected in a population-level association. Therefore, the lack of association is unlikely to be explained by inadequate power, although very small associations cannot be entirely excluded.

### 4.2 Interpreting the null in the context of internal dose

A plausible explanation for the null association is that internal acrylamide doses in the general U.S. population fall well below the thresholds for clinically detectable nerve injury (Fig. 4). In our sample, HbAA and HbGA concentrations (tens of pmol/g globin) were comparable to previously reported reference values (Bergmark, 1997) but remained one to two orders of magnitude below occupational benchmarks. Specifically, these levels are significantly lower than the symptom threshold reported in tunnel workers (∼1,000 pmol/g), the estimated NOAEL (∼2,000 pmol/g), and the LOAEL for peripheral damage (∼6,000 pmol/g) (Bergmark, 1997; Calleman et al., 1994; Hagmar et al., 2001). Given these benchmarks, a null association is the expected outcome predicted by the dose–response literature.

**Figure 4.**
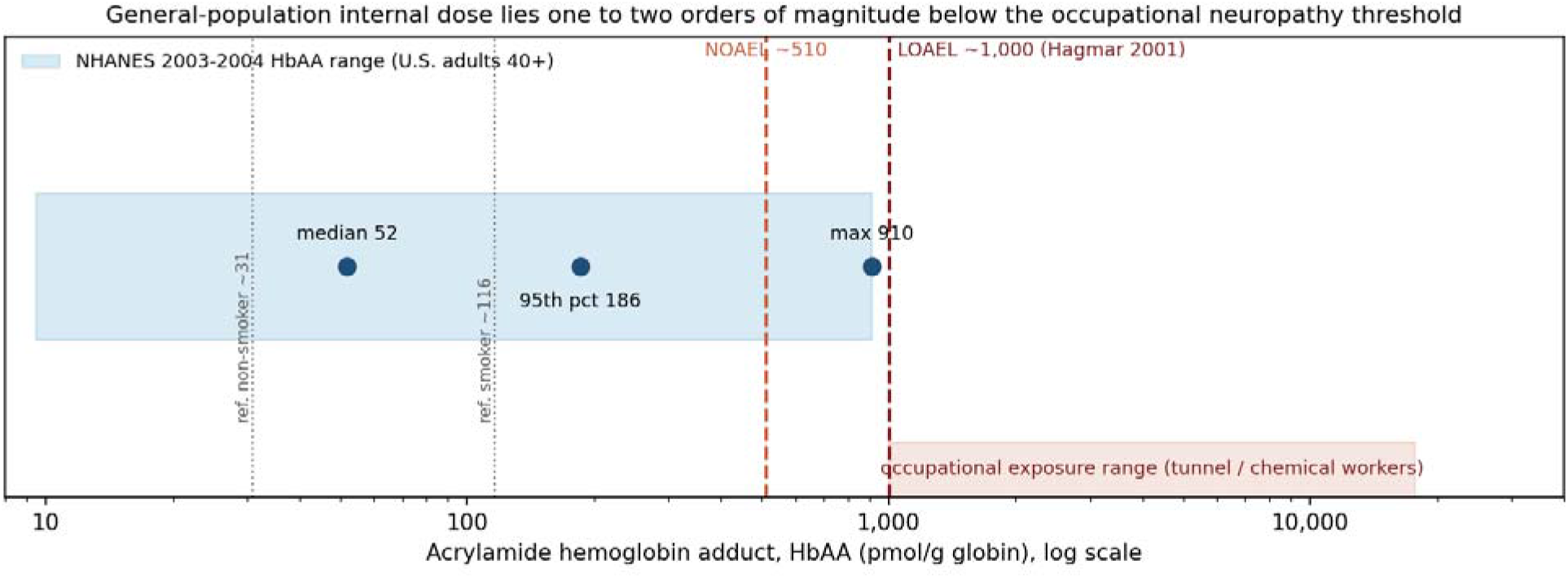
Comparison of acrylamide biomarker levels in the general U.S. population with published occupational exposure benchmarks for neuropathy. Population HbAA and HbGA concentrations were substantially lower than internal-dose thresholds associated with symptom onset, NOAEL, and LOAEL estimates, supporting the observed null association between acrylamide exposure and peripheral neuropathy.

This interpretation is reinforced by the dose-rate dependence of acrylamide neurotoxicity. Experimental work demonstrates that slow, low-level exposure produces qualitatively milder pathology than the same cumulative dose delivered rapidly (Crofton et al., 1996; LoPachin et al., 2000). The chronic, low-flux dietary exposure typical of the general population is unlikely to overwhelm axonal transport systems or exhaust the cellular repair and protein-turnover mechanisms that offset low-level adduction. Consequently, both the low absolute internal dose and the slow kinetics of exposure in the general population argue against detectable large-fiber neuropathy, a conclusion concordant with our results

### 4.3 Concordance with the population biomarker literature

Our results align with evidence that general-population acrylamide doses are inconsistently associated with non-cancer outcomes. Prior NHANES analyses reported null or inverse associations with metabolic syndrome (Hung et al., 2021), chronic kidney disease (Xu et al., 2025), and insulin resistance (Lin et al., 2009). Although dietary acrylamide has been linked to mild cognitive decline in elderly non-smokers (Liu et al., 2017), that central-nervous-system outcome differs fundamentally from the peripheral sensory function assessed here. Moreover, acrylamide does not accumulate significantly even in end-stage renal disease, implying that standard kidney function variation is unlikely to generate the internal exposures required for neurotoxicity (Bahner et al., 2004). This study extends the existing literature to an objectively measured neurological endpoint.

### 4.4 Biological plausibility of a true null

Several factors support the biological plausibility of a null association. First, hemoglobin adducts index systemic exposure but are proxies rather than direct measures of the dose delivered to peripheral axons. Second, humans metabolize acrylamide into glycidamide less efficiently than rodents, and glutathione conjugation further reduces the reactive burden compared to experimental models (Behrens et al., 2019). Third, the high correlation between HbAA and HbGA (r≈0.84) confirms that both adducts index the same exposure pathway; the lack of a signal for either—individually or after mutual adjustment—is therefore internally coherent. Finally, animal studies typically employ doses that vastly exceed human dietary levels, leading to overestimates of risk when extrapolated to population-level exposures (LoPachin, 2005).

### 4.5 Strengths

This study has several methodological strengths. First, the large, nationally representative sample ensures high generalizability to the non-institutionalized U.S. population. Second, both exposure and outcome were assessed objectively—via standardized reference-laboratory biomarkers and clinical examinations—eliminating the recall and reporting biases common in self-reported data. Smoking, a primary non-dietary source of acrylamide, was rigorously controlled using both questionnaire data and objective serum cotinine levels.

Finally, the statistical framework was exceptionally robust. We utilized survey-weighted Taylor-series linearization and subjected our results to an extensive panel of sensitivity analyses. The inclusion of E-values, explicit benchmarking against occupational thresholds, and a formal power analysis collectively demonstrate that our null result is informative and not an artifact of low statistical precision.

### 4.6 Limitations

Several limitations warrant consideration. The cross-sectional design precludes causal inference, though reverse causation is unlikely. Furthermore, while hemoglobin adducts integrate exposure over months, they may not reflect the decades of cumulative dose relevant to chronic axonopathy. Regarding the outcome, monofilament testing identifies manifest large-fiber sensory loss but may miss subclinical or small-fiber neuropathy; such misclassification would likely bias estimates toward the null.

Generalizability is limited by the age restriction (≥40 years) and the use of a single survey cycle (2003–2004). However, these data remain the unique national benchmark pairing internal biomarkers with objective clinical examinations. Finally, a population-average null result does not exclude risks within highly susceptible subgroups—such as individuals with high CYP2E1 activity or GST-null genotypes—who could not be resolved at the current study’s resolution.

### 4.7 Implications and future directions

For the general population, these findings are reassuring regarding clinically detectable large-fiber neuropathy at contemporary exposure levels. This result does not, however, diminish regulatory concerns regarding acrylamide’s genotoxicity and carcinogenicity, nor does it exclude potential developmental neurotoxicity or effects within high-exposure occupational groups.

Future work should employ prospective cohorts with repeated adduct measurements to capture long-term cumulative dose. Research should also utilize more sensitive neurological phenotyping, including quantitative sensory testing and small-fiber assessment. Evaluating gene– environment interactions involving metabolic polymorphisms remains a priority for identifying potentially vulnerable subpopulations.

## 5. Conclusion

In a nationally representative sample of U.S. adults with biomarker-confirmed internal dose and an objective neurological outcome, acrylamide and glycidamide hemoglobin adducts were not associated with peripheral neuropathy. These findings are consistent with the hypothesis that population internal doses remain below levels associated with clinically detectable peripheral neuropathy in occupational settings, and is reinforced by the demonstrated ability of the same models to detect the expected effects of age, sex, and diabetes. At contemporary levels of predominantly dietary exposure, these data do not support an association between acrylamide biomarkers and clinically detectable large-fibre peripheral neuropathy in the general adult population, while its carcinogenic and developmental hazards and possible subclinical or subgroup-specific effects remain priorities for continued investigation.

## Author Contributions

R.M.B., N.N., M.A.S., R.S.M.T., M.G. and K.H. contributed to study conception and design. K.H. performed data analysis and drafted the manuscript. All authors reviewed, revised, and approved the final manuscript.

## Funding

This research received no external funding.

## Data Availability Statement

The data used in this study are publicly available from the National Health and Nutrition Examination Survey (NHANES) database (https://www.cdc.gov/nchs/nhanes/about/index.html).

## Declaration of Competing Interest

The authors declare that they have no known competing financial interests or personal relationships that could have appeared to influence the work reported in this paper.

## Acknowledgments

The authors thank the participants and staff of the National Health and Nutrition Examination Survey (NHANES) for making these data publicly available.

## Declaration of Generative AI and AI-assisted Technologies in the Writing Process

During the preparation of this manuscript, the authors used Gemini 3 as an AI-assisted writing tool to support language refinement, text editing, formatting, and improvement of manuscript readability. The authors carefully reviewed, revised, and validated all generated content and take full responsibility for the accuracy, integrity, and originality of the final manuscript. The AI tool was used solely to assist in the writing process and did not contribute to the scientific conclusions, data analysis, interpretation of results, or authorship of the work.

## Ethics Approval and Consent to Participate

This study is a secondary analysis of publicly available, de-identified data from the National Health and Nutrition Examination Survey (NHANES). NHANES protocols and procedures were approved by the National Center for Health Statistics (NCHS) Research Ethics Review Board (ERB) in accordance with the ethical principles of the Declaration of Helsinki. All human participants provided written informed consent prior to participation

## Clinical trial number

Not applicable.

---

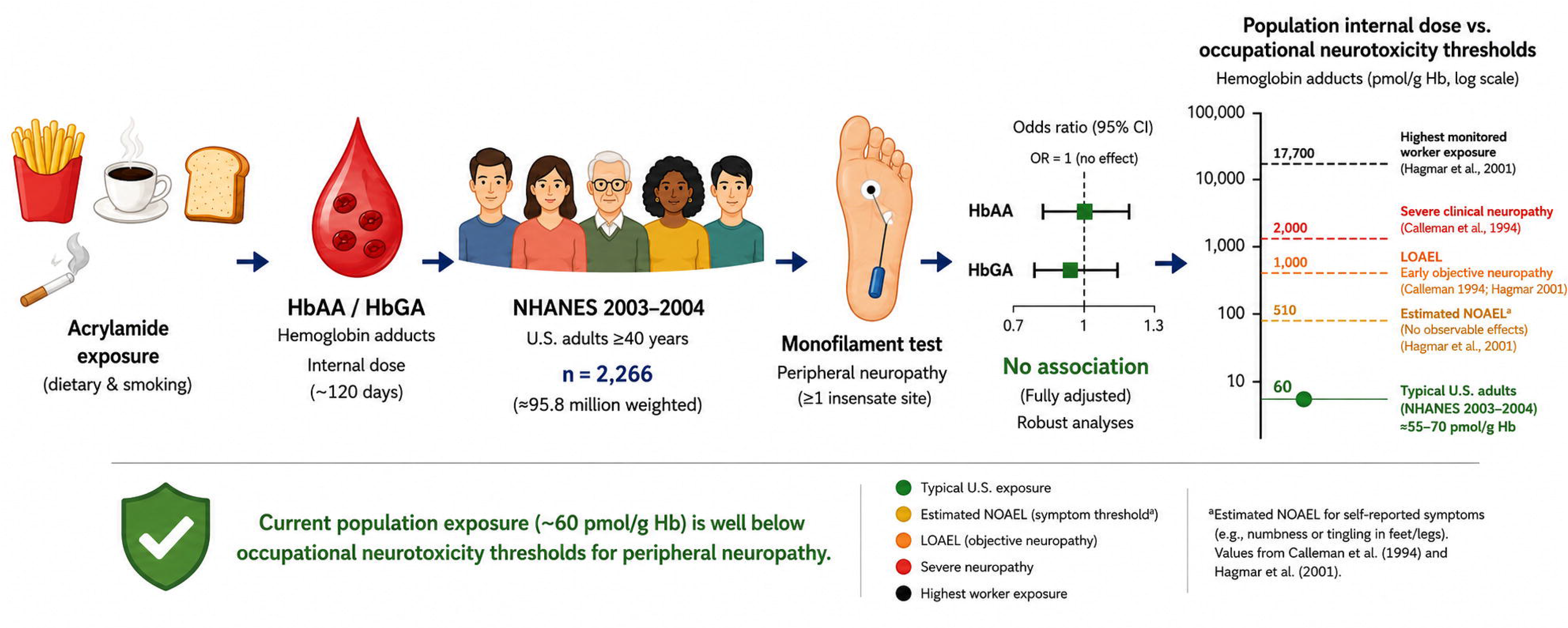

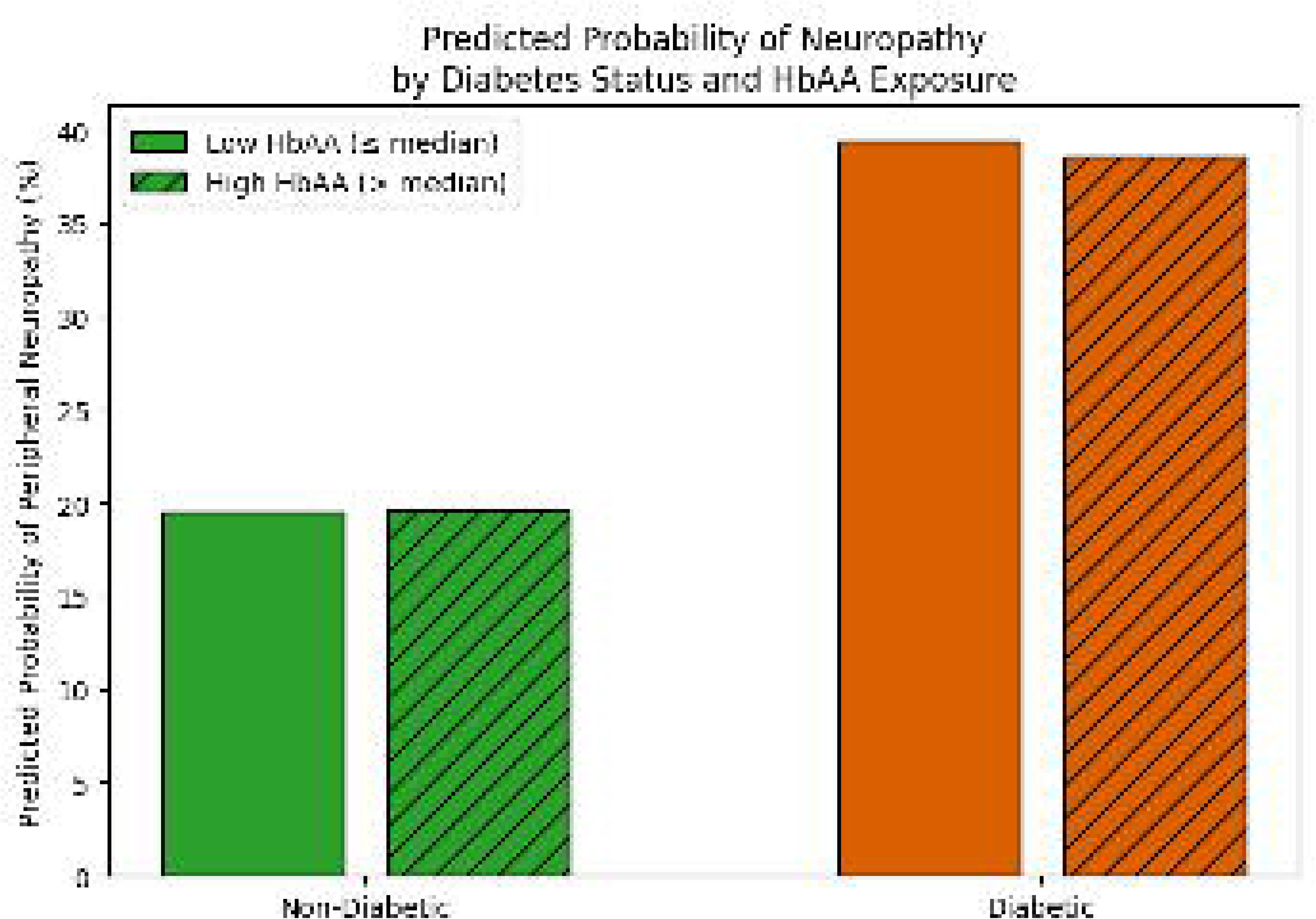

## Notes

### Competing Interest Statement

The authors have declared no competing interest.

